# Post-Ablation PAI-1 Difference as a Biomarker for Ablation Success: A Diagnostic Study

**DOI:** 10.1101/2025.10.18.25338273

**Authors:** Roin Rekvava, Jay Nemade, Teimurazi Tchikhoria, Salim Berkinbayev, Nino Tabagari

## Abstract

**Background:** Atrial fibrillation (AF) is the most common sustained cardiac arrhythmia and is associated with a prothrombotic state and impaired fibrinolysis. Plasminogen activator inhibitor-1 (PAI-1), a key regulator of the fibrinolytic system, is linked to increased cardiovascular risk when elevated. Catheter ablation is a well-established treatment for restoring sinus rhythm in AF. Previous studies show that PAI-1 levels decrease 1–3 months post-ablation, indicating potential therapeutic relevance.

**Objective:** To evaluate whether within-patient differences in PAI-1 levels predict procedural success following catheter ablation for AF.

**Methods:** 70 AF patients aged ≥60 years (35 paroxysmal, 35 persistent) undergoing ablation were prospectively enrolled. PAI-1 levels were measured pre-procedure and 1–3 months post-ablation. Differences were correlated with ablation modality, AF type, recurrence and sex. Analyses included Spearman’s correlation, ROC curves, and diagnostic performance metrics. Difference thresholds were determined using interpolation, Highest-Density-Interval, and Bayesian analysis.

**Results:** Overall, PAI-1 levels decreased significantly post-ablation (t=15.1, p<0.00001); weak to moderate differences were observed by sex (t=2.14, p<0.05) and AF type (t=4.26, p<0.0001). Post-ablation PAI-1 difference (–14 to –3.48 ng/mL) predicted a recurrence rate of 4.2–22.5%, with 92% sensitivity, 65% specificity, 87% PPV, 76% NPV, and 84% accuracy.

**Conclusion:** In patients aged ≥60 years, within-patient PAI-1 differences of –14 to –3.48 ng/mL may help predict ablation outcomes. Although limited by small sample size, mechanistic rationale suggests that greater reductions in PAI-1 may be associated with improved procedural performance.

## Introduction

### Background

Atrial fibrillation (AF), the most common sustained arrhythmia, creates a prothrombotic state through endothelial dysfunction, platelet activation, and impaired fibrinolysis, increasing stroke risk. A key mechanism involves dysregulation of fibrinolysis: Tissue Plasminogen Activator (t-PA) promotes clot breakdown, while its inhibitor Plasminogen Activator Inhibitor-1 (PAI-1) suppresses this process [1]. Elevated PAI-1 reflects inflammation, contributes to thrombosis, and is closely linked to AF pathophysiology [2]. Paroxysmal AF, though transient, carries a thromboembolic risk similar to persistent AF [3].

### Indications for the study

A prospective study at Loyola University Medical Center evaluated 53 patients with AF undergoing catheter ablation, measuring circulating PAI-1 and CD40 ligand levels. Compared with healthy controls, baseline levels of both markers were elevated. Following ablation, CD40 ligand levels decreased significantly at 1 and 3 months, while PAI-1 levels declined at 1 month. These findings support PAI-1 as a potential biomarker of thromboembolic risk and procedural success, and suggest that ablation favorably modulates inflammatory and fibrinolytic pathways [4].

A prospective study at Vanderbilt University analyzed 253 patients undergoing cardiac surgery with cardiopulmonary bypass, 26.5% of whom developed postoperative AF, peaking on day 2. Higher PAI-1 levels (both pre- and postoperative) were strongly associated with AF, alongside age and prior AF history. Multivariate analysis confirmed preoperative PAI-1 as the strongest predictor. These findings suggest that elevated PAI-1 contributes to postoperative AF and that modulating PAI-1 may represent a viable therapeutic strategy [5].

The Catheter Ablation vs Antiarrhythmic Drug Therapy for Atrial Fibrillation (CABANA) trial randomized 2,204 patients with AF to catheter ablation or medical therapy. Ablation significantly reduced AF recurrence (49.9% vs. 69.5%; HR 0.52; *P* < 0.001) and the composite of death or cardiovascular hospitalization (51.7% vs. 58.1%; HR 0.83; *P* = 0.001). However, no significant differences were observed in all-cause mortality or stroke, underscoring the need for careful patient selection and balanced consideration of rhythm-control benefits [6].

The FIRE AND ICE trial randomized 762 patients with drug-refractory paroxysmal AF to cryoablation or radiofrequency ablation (RFA). At one year, cryoablation was non-inferior to RFA (event rates: 34.6% vs. 35.9%; HR 0.96; *P* < 0.001) with a comparable safety profile. Cryoablation offered potentially shorter procedure times and less thrombotic activation, supporting its role as an effective and safe alternative to RFA [7].

### Research Gap & Rationale

Previous studies [4-7] have examined dynamic biomarkers to predict ablation success, emphasizing clinical outcomes and procedural efficacy. Genetic, environmental, pathological, and physiological factors have significant influences on PAI-1 levels [8]. Given these influences, we find that within-subject comparisons (especially in defined subgroups), are likely more reliable than baseline levels alone. Also, serial measurement of PAI-1 levels before and after ablation reflects thrombotic and fibrinolytic balance, filling gaps in existing research and offering a practical tool for outcome prediction. This strategy may enable more personalized patient care and improve assessment of ablation efficacy.

Thus, in this study we verify earlier studies while exploring within-patient PAI-1 level differences as a post-ablative success metric.

## Methodology

### Study population

A total of 70 patients with AF were enrolled according to predefined inclusion criteria. Baseline demographic and clinical data were collected at enrollment. Potential confounders, including comorbidities, inflammatory status, and thrombotic profile, were not controlled, as the primary goal of this study was to analyze within-patient PAI-1 differences as a biomarker. These factors are acknowledged as limitations that may influence interpretation of the results.

Given the exploratory nature of the study, no formal sample size calculation was performed.

### Inclusion and Exclusion Criteria

The study cohort included 35 patients with paroxysmal AF and 35 with persistent AF. Patients were randomly assigned to either radiofrequency ablation or cryoablation to balance known and unknown confounders, including age, AF type, anticoagulation intensity, inflammatory status, and operator skill. Patients with a left atrial diameter outside the range of 40–52 mm were excluded to ensure atrial size homogeneity. Eligible participants were also required to have a documented AF history of 2–10 years, to minimize confounding from early-stage disease, while avoiding heterogeneity associated with long-standing AF and advanced atrial remodeling.

### Measurement of PAI-1 Levels

Baseline PAI-1 levels were obtained approximately two weeks prior to ablation using ELISA (Enzyme-Linked Immunosorbent Assay). Follow-up sampling was performed 1–3 months post-ablation for most patients; in two cases, logistical challenges delayed sampling to 6 months. This variability in follow-up timing is consistent with the approach used by [9], which collected post-ablation samples within a 1–3-month window to capture PAI-1–related changes while allowing flexibility in post-procedural sampling.

### Follow-up and Outcome Definition

Patients were intended to be monitored for cardiac rhythm status for up to 12 months after ablation. Rhythm surveillance was performed through scheduled clinic visits, including clinical evaluation and documentation of arrhythmic symptoms or ECG confirmed AF episodes. Follow-up was discontinued for patients who experienced recurrence or when continued monitoring was deemed unnecessary to avoid imposing additional burden on the patient. Any documented recurrence of AF during follow-up classified the intervention as non-successful. Patients without AF recurrence at their last available follow-up visit were considered recurrence-free.

Consistent with previous studies [10–12], this binary classification of patients as either experiencing recurrence or remaining recurrence-free is well-supported and was adopted as the primary clinical outcome.

### Data Analysis

A preliminary analysis was conducted in the context of previously published literature to establish baseline comparability and identify potential trends. Normality of age, AF Duration and LA Diameter was checked using Shapiro-Wilk test. Differences in PAI-1 levels (pre-vs. post-ablation) were graphically represented, with color coding to stratify patients according to clinical and procedural parameters, enabling identification of differentiating trends across strata. Spearman’s correlation, Chi-squared test, Mann–Whitney U tests, independent t-test and Wilcoxon signed-rank tests were used to evaluate associations and differences of PAI-1 levels across ablation modality, AF type, sex, and recurrence status. All statistical analyses and tests were performed using Python 3.11.1 with NumPy, SciPy, pandas, and scikit-learn; visualizations were generated using bokeh. Two-tailed p-values <0.05 were considered statistically significant.

### Establishing Laboratory Reference Ranges for PAI-1

Kernel Density Estimation (KDE) [13] has been shown to effectively interpolate data ranges; therefore, it was applied here to address the dataset’s small sample size. KDE was used to model PAI-1 differences and generate Probability Density Functions (PDFs) for the overall, recurrent, and non-recurrent groups. Thresholds were derived from the Highest Density Interval (HDI), procedural success estimated using Bayes’ theorem, and predictive performance evaluated with standard diagnostic metrics.

Receiver operating characteristic (ROC) curves were generated, and area under the curve (AUC) values calculated to assess discriminative ability. Sensitivity, specificity, and related indices were computed to quantify accuracy and potential clinical applicability, with 95% confidence intervals. Additional markers [14], such as left atrial diameter, were also analyzed for comparison.

### Ethics

All participants provided written informed consent prior to enrollment, including consent for collection and analysis of clinical data and biological samples. The study protocol was deemed exempt from formal ethical approval under local regulations governing observational biomarker research (Personal Data Protection Service, Georgia). The study was conducted in accordance with the Declaration of Helsinki. Patient data were fully anonymized, and written consents are retained by the investigators and available upon request.

## Results

**Data Characteristics and Demographics**

Patients had a mean age of 69.6 ± 5.9 years, median 70 (IQR 64.3–74.8), and age ranged from 60 to 81 years. Mean AF duration was 5.5 ± 2.4 years, median 5.5 (IQR 4–7), with a range of 2–10 years. The mean left atrial diameter was 45.8 ± 3.4 mm, median 45 (IQR 43–48.8). In paroxysmal AF, patients were older (mean 74.4 ± 3.3 years) with longer AF duration (7.4 ± 1.6 years) and smaller atria (43.1 ± 1.8 mm) compared to persistent AF (mean age 64.6 ± 3.4 years, AF duration 3.6 ± 1.3 years, LA diameter 48.5 ± 2.3 mm). Sex distribution was 41.4% female, 58.6% male, and ablation modality included RFA in 49 and cryoablation in 21 patients. Shapiro–Wilk testing indicated non-normal distributions for age, AF duration, and LA diameter (all p < 0.05). Observed follow-up ranged from 3 to 10 months, with both mean and median follow-up of 6 months. All patients received anticoagulation therapy, with the majority on direct oral anticoagulants and 8 managed with Warfarin for clinical indications.

### Baseline PAI-1 levels and post-ablation differences

In our cohort, pre-ablation PAI-1 levels had a mean of 32.6 ng/mL ± 5.3 ng/mL (range 22.5–42.5 ng/mL) and a median of 34.4 ng/mL (IQR 29.1–36.2 ng/mL). Following ablation, PAI-1 levels decreased in nearly all patients, with changes ranging from –21.8 to -0.8 ng/mL; only one patient exhibited a slight increase of 1.4 ng/mL. The reduction was statistically significant (median change–9.8 ng/mL, Wilcoxon signed-rank test W = 0, p < 0.0001).

Spearman’s correlation demonstrated a strong positive association between pre- and post-ablation levels (ρ = 0.85, p < 0.0001). No significant correlation was observed between baseline PAI-1 levels and the magnitude of post-ablation change (Spearman’s ρ = 0.09, p = 0.45).

Females exhibited a mean PAI-1 change of –7.7 ± 4.4 ng/mL (Mdn –10.3 ng/mL, IQR –11.5 to – 3.5 ng/mL), while males showed a mean change of –7.9 ± 4.3 ng/mL (Mdn –8.5 ng/mL, IQR –10.7 to –5.0 ng/mL). Paroxysmal AF patients observed a mean change of –7.6 ± 4.2 ng/mL (Mdn –10.4 ng/mL, IQR –11.4 to –3.5 ng/mL), and persistent AF patients a mean change of –8.0 ± 4.5 ng/ mL (Mdn –9.6 ng/mL, IQR –10.7 to –7.0 ng/mL).

Scatter plots of pre-versus post-ablation PAI-1 levels, color-coded by sex and AF type, revealed no distinct separation or trend between groups (Figures 1.A and 1.B). These observations were supported statistically by Mann–Whitney U tests, showing no significant differences in PAI-1 changes between sexes (U = 669.5, p = 0.374) or AF types (U = 693.0, p = 0.347).

**Figure 1.**
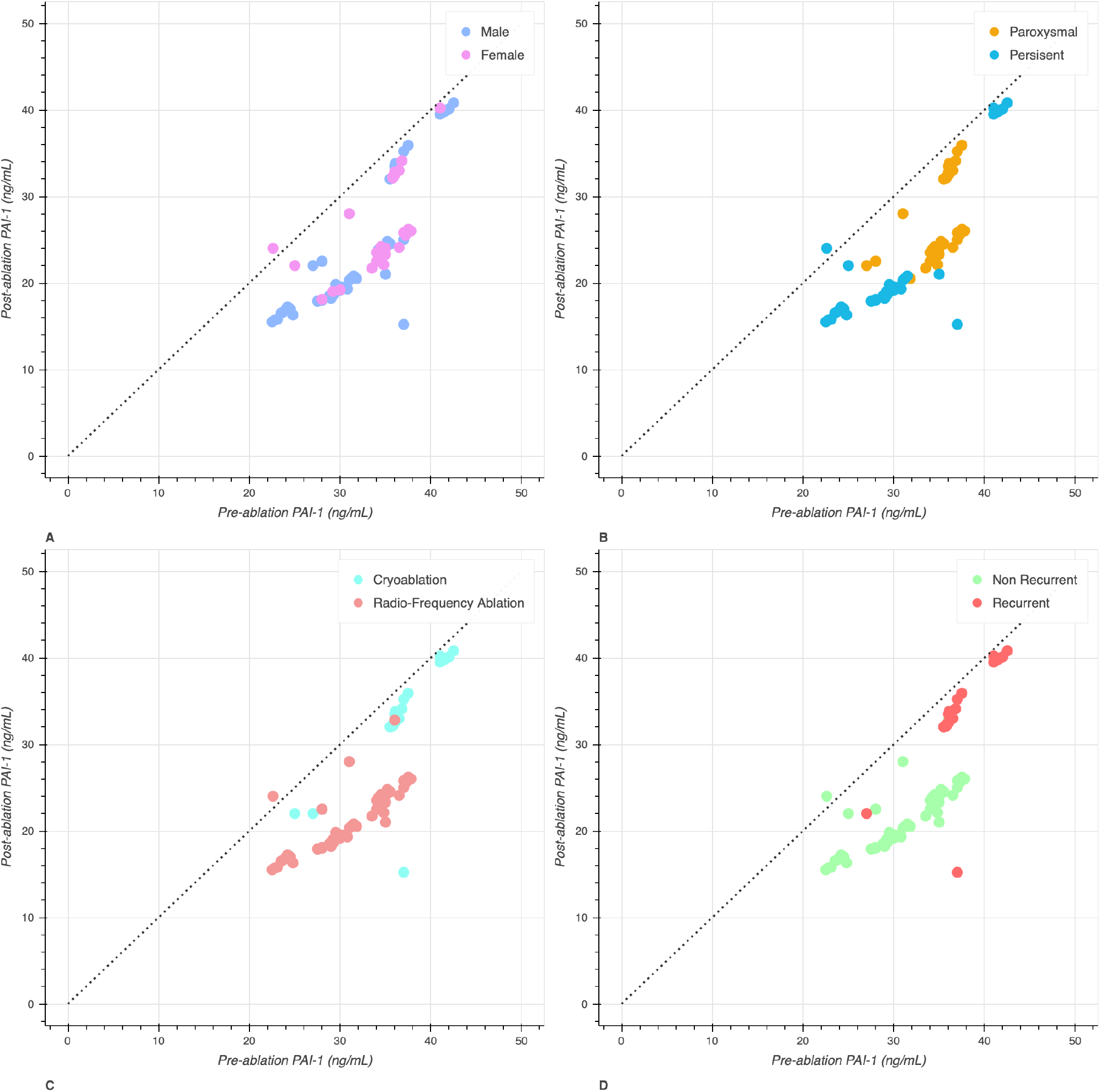
Scatter plots of pre- versus post-ablation PAI-1 levels: Each point represents an individual patient. Plots are color-coded by sex (A), AF type (B), ablation modality (C), and recurrence status (D). The dashed line represents equality, indicating no change between pre- and post-ablation PAI-1 levels.

While normality testing indicated significant deviations in both groups (Shapiro–Wilk, p < 0.00001), visual inspection of the KDE probability density functions suggested approximately skewed, leptokurtic bell-shaped distributions. Recurrent patients exhibited a mean PAI-1 change of –3.6 ± 4.4 ng/mL (Mdn –2.6 ng/mL, IQR –3.5 to –1.8 ng/mL), whereas non-recurrent patients showed a mean change of –9.5 ± 2.9 ng/mL (Mdn –10.5 ng/mL, IQR –11.3 to –7.8 ng/mL). This difference was statistically robust (U = 907.0; t = 6.586; p < 0.00001).

Similarly, cryoablation patients demonstrated a mean change of –3.6 ± 4.3 ng/mL (Mdn –2.7 ng/ mL, IQR –3.5 to –1.8 ng/mL), compared with –9.6 ± 2.8 ng/mL (Mdn –10.5 ng/mL, IQR –11.3 to – 8.5 ng/mL) in the RFA group. This difference was highly significant (U = 943.5; t = 5.07; p < 0.00001). Scatter plots of pre-versus post-ablation PAI-1 levels (Figures 1.C and 1.D) showed a 95.2% overlap between cryoablation and recurrent patients, while also revealing a clear separation in trends between cryoablation and RFA. Consistent with these findings, cryoablation was strongly associated with recurrence (χ^2^(1) = 60.75, p < 0.0001).

### PAI-1-Based Analysis of AF Recurrence

The recurrence of the overall study population was 28.5%. Stratified by subgroups, recurrence occurred in 20.0% of persistent AF patients, 37.1% of paroxysmal AF patients, 0% of patients treated with RFA, and 95.2% of those treated with cryoablation. Corresponding success rates were 80.0%, 62.9%, 100%, and 4.8%, respectively.

In the KDE-interpolated PDFs (Figure 2), recurrent patients exhibited a negatively skewed, highly peaked distribution (skewness = –4.07, kurtosis = 13.12), whereas non-recurrent patients showed a positively skewed, moderately peaked distribution (skewness = 1.72, kurtosis = 2.9). The 95% HDI for post-ablation PAI-1 differences in the successful group ranged from –14.0 to –3.48 ng/mL, encompassing 91% of successful and 35% of recurrent patients. Applying Bayes’ theorem to our cohort (N = 50 non-recurrent, R = 20 recurrent) yielded a recurrence probability of 13.3% within this range (95% CI: 4.2–22.5%).

**Figure 2.**
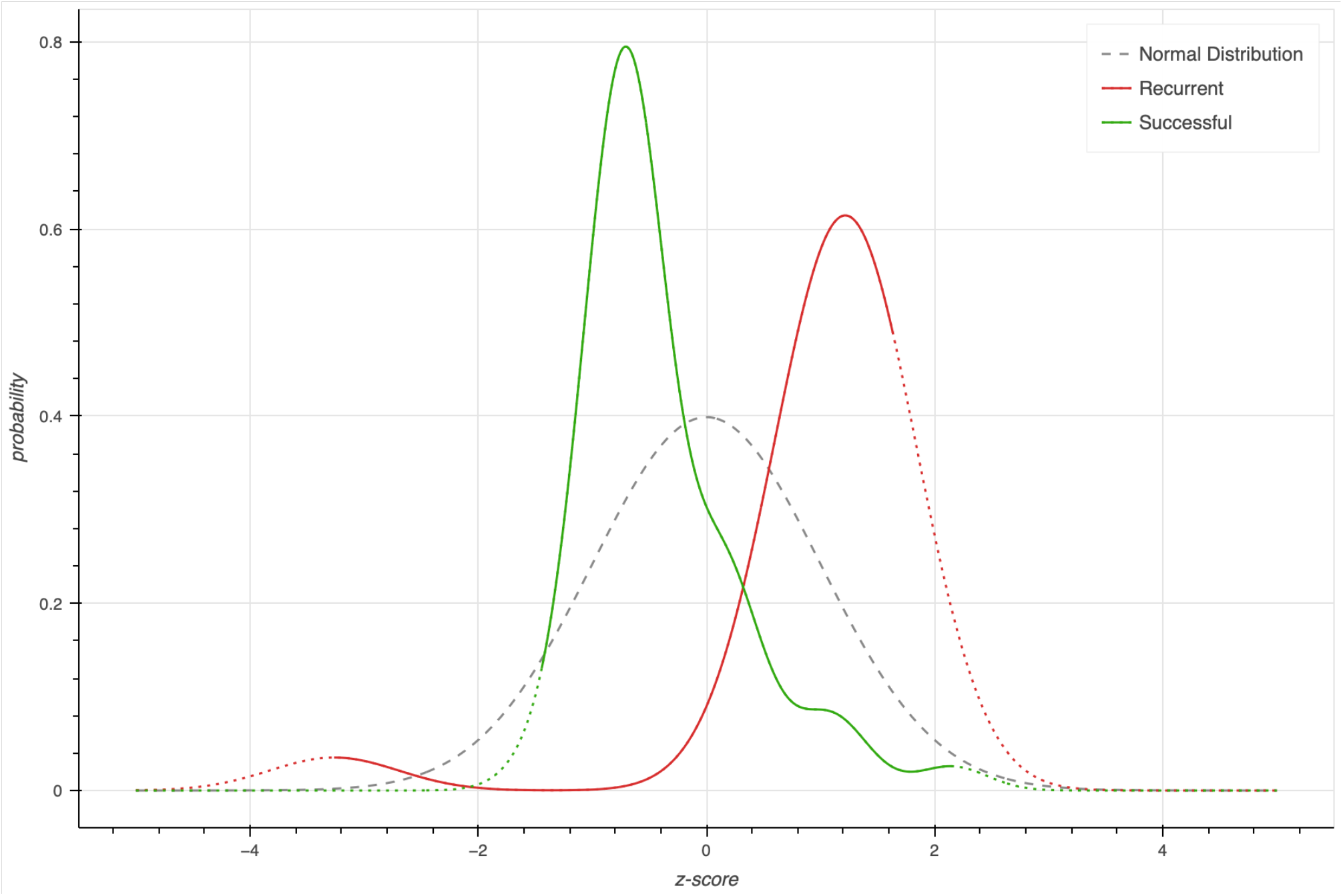
KDE derived PDFs of recurrent and successful patients: The graph shows z-score vs probability of PAI-1 differences, where the dotted lines show extended data, and the bold lines show actual data range of our cohort. A normal distribution is also plotted for reference.

### Diagnostic Metrics

ROC analyses (Figure 3) evaluated predictors of AF recurrence. LA diameter demonstrated poor discrimination (AUC = 0.486), while PAI-1 difference showed good performance (AUC = 0.907). Applying the –14.0 to –3.48 ng/mL reference range achieved perfect discrimination (AUC = 1.0).

**Figure 3.**
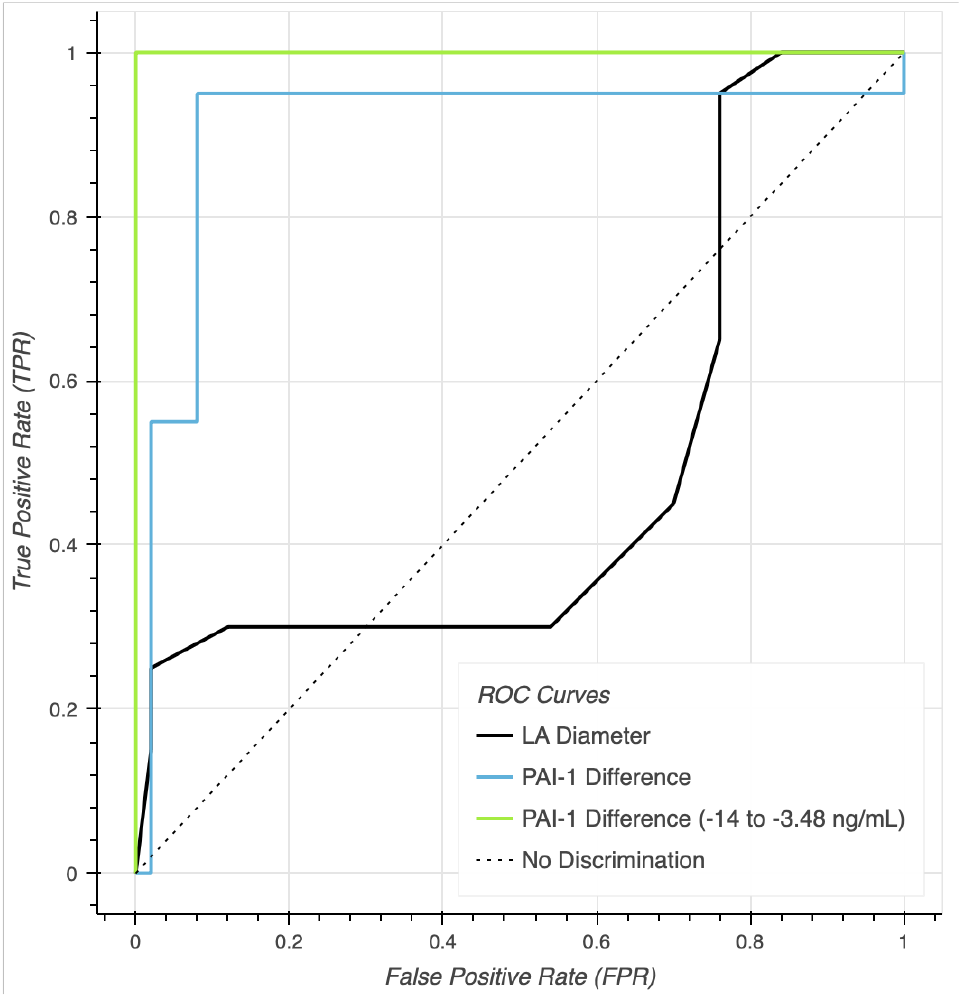
ROC Analyses.

Contingency table analysis (Table 2) demonstrated that using the test range of –14 to –3.48 ng/mL yielded a sensitivity of 92%, specificity of 65%, positive predictive value (PPV) of 86.8%, negative predictive value (NPV) of 76.5%, and an overall accuracy of 84.3%.

**Table 1.**
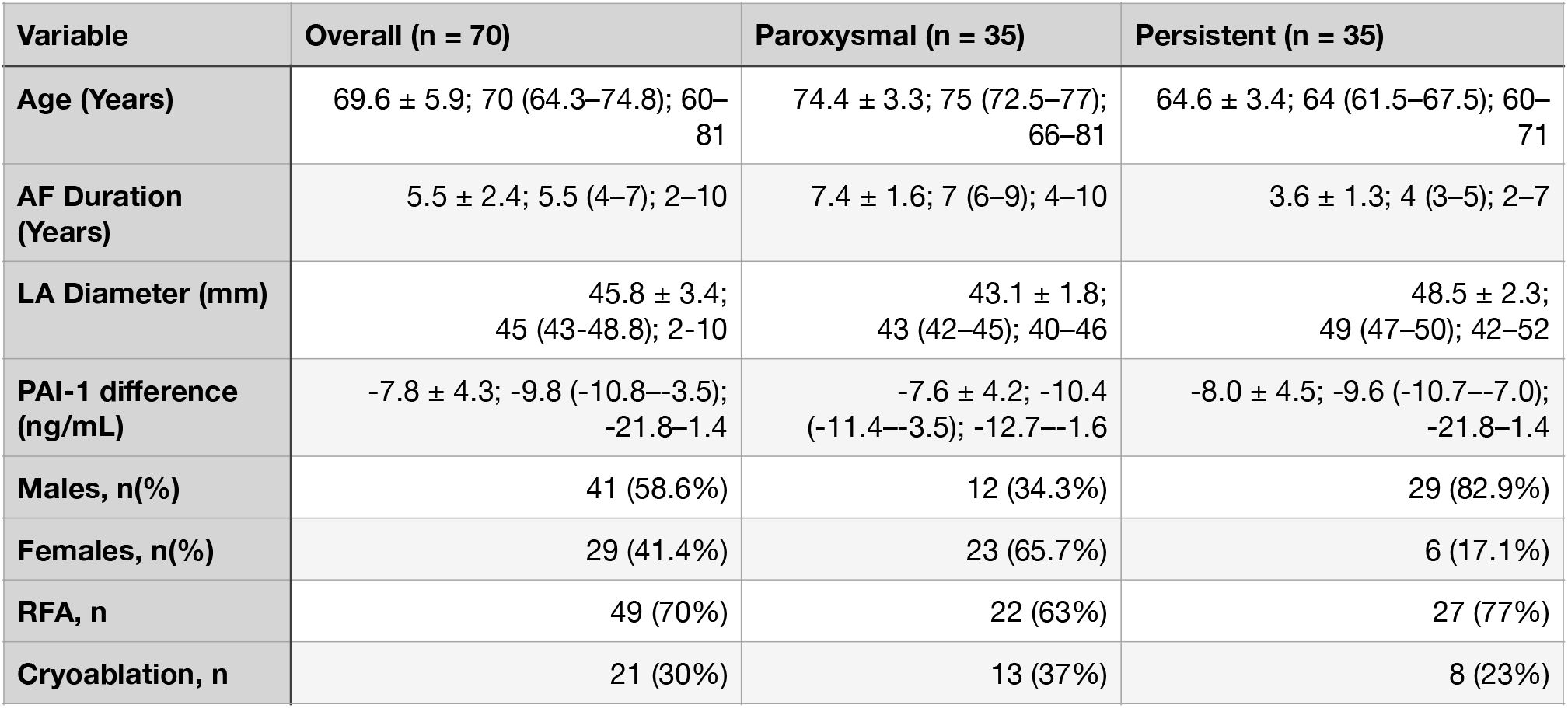
Baseline Characteristics of the Cohort and by AF Type: Values are presented as mean ± standard deviation, median (IQR), and minimum–maximum for continuous variables. Categorical variables are shown as n (%).

**Table 2.**
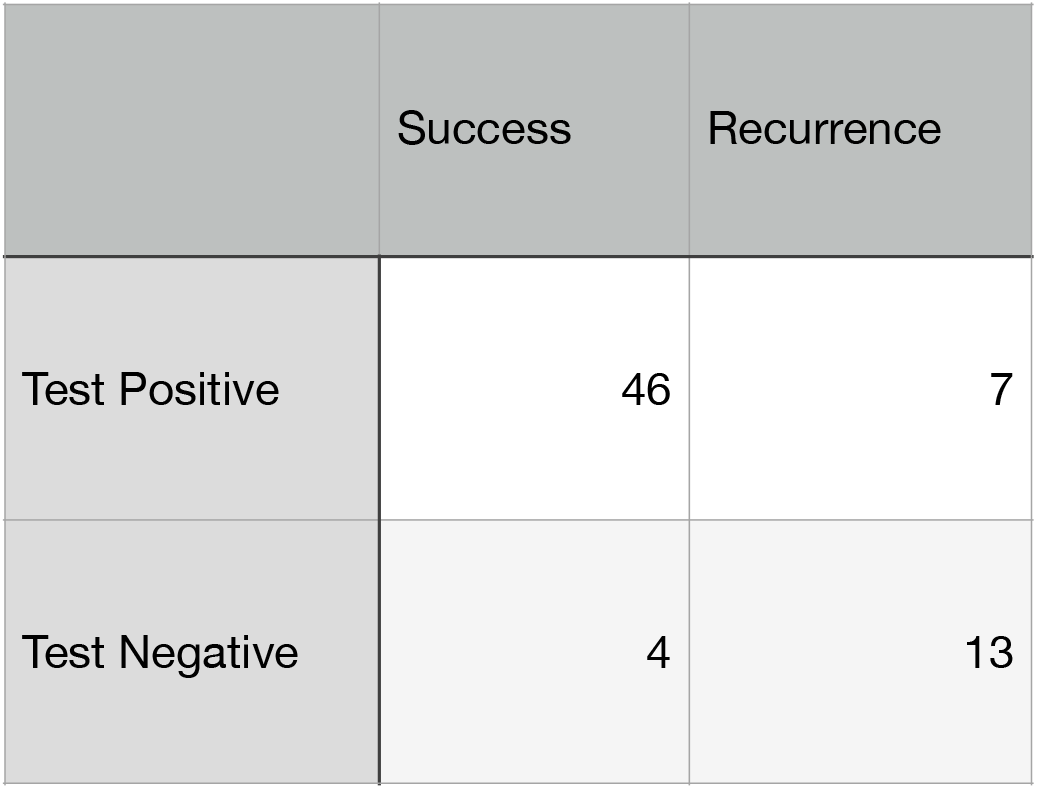
Contingency Table.

## Discussion

### Principal Findings

The observed non-normality is unsurprising, given that the cohort comprises elderly AF patients (≥60 years, mean age ∼70 years) with mildly enlarged left atria and an average AF duration of ∼4 years. The modest male predominance (17.2%) aligns with typical sex distributions in this population. Overall, these characteristics indicate that our cohort is representative of a typical elderly AF population, consistent with our other findings.

In our cohort, PAI-1 levels consistently decreased (Mdn –9.8 ng/mL, range –21.8 ng/mL to 1.4 ng/ mL) following ablation, supporting the notion that the procedure modulates fibrinolytic and thrombotic pathways. Although pre- and post-ablation levels were strongly correlated (p < 0.0001), the magnitude of change was independent of baseline values (p > 0.05), indicating that within-patient PAI-1 difference is robust to individual variability and a reliable marker of procedural success.

No significant differences were observed between AF types or sexes (both p > 0.05), thereby excluding these variables as potential confounders of post-ablation PAI-1 change. Similarly, subgroup analyses revealed no significant within-group variation (p > 0.05). Collectively, these findings suggest that PAI-1 reduction is largely similar across sex and AF form, reinforcing its validity as a biomarker in the broader AF population.

By contrast, the post-ablation difference between recurrent and non-recurrent groups was highly significant (p < 0.0001), indicating that PAI-1 change is a strong discriminator of recurrence. ROC analysis confirmed this, yielding an AUC of 0.907, corresponding to ∼91% discrimination between recurrent and non-recurrent patients.

A highly significant (p < 0.0001) difference was also observed between RFA and cryoablation, with separate trends suggesting cryoablation reduces PAI-1 levels post-ablation less than RFA. Consistent with these findings, cryoablation was strongly associated with recurrence (p < 0.0001).

Using these data, we established a prospective post-ablation reference range of –14 to –3.48 ng/ mL, which demonstrated a sensitivity of 92%, specificity of 65%, PPV of 86.8%, NPV of 76.5%, and overall accuracy of 84.3%. Within this range, the estimated procedural success rate is approximately 87%, though given the characteristics of our cohort, the true rate may plausibly range from 77.5% to 95.5%.

### Comparison with Existing Literature

Our finding that post-ablation PAI-1 differences are significantly greater in the recurrent group than in the non-recurrent group (p < 0.0001) aligns with the Loyola study, which reported elevated baseline PAI-1 levels in AF patients compared with controls (19.55 ± 2.17 ng/mL vs. 4.85 ± 0.41 ng/mL, P < 0.0001).

Likewise, the modestly enlarged left atrial diameters in our cohort are consistent with the Vanderbilt study. Building on these observations, our results show consistent post-ablation reductions in PAI-1, with greater decreases following RFA compared to cryoablation, correlating with better clinical outcomes. Together, these findings reinforce the concept that modulating PAI-1 levels represents a viable therapeutic strategy to improve ablation success, as previously suggested by Vanderbilt investigators.

The observed recurrence rate of 28.5% in our cohort is consistent with the CABANA trial, which established catheter ablation as an effective strategy for AF management, though without clear superiority over medical therapy alone. Notably, our finding of higher recurrence among cryoablation patients highlights a potential gap in CABANA, where procedural differences were not fully addressed. These results suggest that the efficacy of specific ablation techniques may differ and that PAI-1 could serve as a useful biomarker for stratifying outcomes across procedural modalities.

Our findings contrast with the FIRE AND ICE trial [7], which reported non-inferiority of cryoablation versus RFA. In our cohort, cryoablation was linked to recurrence and smaller PAI-1 reductions, while RFA yielded greater decreases. While the contrast in recurrence rates may reflect the low sample size, the trend differences suggests that larger post-ablation PAI-1 reductions may indicate procedural efficacy.

### Clinical Implications

Because post-ablation PAI-1 reduction occurs independently of patient sex or AF type, its applicability as a biomarker is strong, remaining consistent across demographic subgroups. While baseline PAI-1 levels tend to rise with age, the absence of correlation between baseline levels and post-ablation change suggests that age is unlikely to act as a confounder. Accordingly, our findings support the use of PAI-1 reduction as a reliable biomarker in AF populations aged 60 years and older, irrespective of AF form (paroxysmal or persistent).

PAI-1 differences within the range of –14.0 to –3.48 ng/mL demonstrated excellent discrimination between recurrent and non-recurrent groups. While our estimated recurrence probability was 13.3%, the limited sample size broadened the confidence interval to 4.2–22.5%. Accordingly, future patients falling within this reference range should be considered to carry a recurrence risk within this interval, corresponding to an estimated ablation success rate of 77.5–95.5%.

### Strengths

The prospective measurement of a dynamic biomarker (PAI-1) captures real-time changes in thrombotic and fibrinolytic balance, providing clinically relevant insights that static baseline values cannot. Additionally, within-subject comparisons further minimize confounding, as each patient effectively serves as their own control. This approach reduces the impact of inter-individual variability arising from genetic, environmental, or pathological factors, thereby strengthening the reliability of observed associations.

Finally, the statistical approach was tailored to the data. Non-parametric methods appropriately addressed non-normal distributions, ROC analysis yielded clinically interpretable measures of diagnostic performance, and Bayesian probability offered outcome likelihoods with quantified uncertainty. Together, these methodological strengths enhance the internal validity of our findings and support their potential clinical applicability.

### Limitations

While strong, our results are limited by the relatively modest sample size of 70 patients, positioning this work as a pilot study to guide future research. The unequal distribution between the RFA and cryoablation groups further constrains our ability to draw definitive conclusions regarding procedural performance.

Notably, the non-recurrence among RFA patients may reflect the limited sample size rather than a true absence of recurrence. Additionally, the low sample size also influences randomization which could explain the high recurrence we observed in cryoablation group.

In addition, the short and variable follow-up period limits assessment of long-term outcomes. We did not control for all potential confounders, such as medications and comorbidities; however, the prospective design and predominant use of DOAC-based anticoagulation help mitigate these effects. Finally, the wide confidence intervals in some analyses restrict the precision of estimated risks and success rates.

### Future Directions

Future studies should validate these findings in larger, multi-center cohorts to ensure broader applicability. Bench studies investigating the mechanisms behind modality-specific PAI-1 changes could clarify how different ablation techniques influence fibrinolytic and thrombotic balance.

Additionally, direct comparisons of RFA and cryoablation in relation to post-ablation PAI-1 modulation and clinical outcomes are needed to provide more definitive evidence of procedural efficacy. Finally, evaluating the impact of pre- or post-procedure prothrombotic or fibrinolytic pharmacological interventions may help improve ablation success, minimize intra-operative damage, and reduce recurrence rates.

## Conclusion

Post-ablation reductions in PAI-1 were consistent across demographics and strongly predictive of recurrence in AF patients aged ≥60 years. A within-patient difference of –14 to –3.48 ng/mL may identify high procedural success, while smaller reductions signaled recurrence risk. RFA patients exhibited greater post-ablation decreases in PAI-1 than cryoablation patients; however, given the small sample size, these observations should be interpreted cautiously. These findings position PAI-1 modulation as a promising biomarker of ablation success, warranting validation in larger, multi-center cohorts with extended follow-up.

## Data Availability

All data produced in the present study are available upon reasonable request to the authors

## Acknowledgments

✴ We sincerely thank all individuals who contributed to this study, particularly those who provided guidance and insights during manuscript preparation.

## List of Abbreviations

AF: Atrial Fibrillation
AUC: Area Under the Curve
DOAC: Direct Oral Anticoagulant
ECG: Electrocardiogram
HDI: Highest Density Interval
KDE: Kernel Density Estimation
PAI-1: Plasminogen Activator Inhibitor-1
RFA: Radiofrequency Ablation
ROC: Receiver Operating Characteristic
t-PA: Tissue Plasminogen Activator

